# *Poisoned in Their Homes* - Red Blood Cell Abnormalities in Lead-Exposed Residents of a Pakistani Industrial Zone

**DOI:** 10.1101/2023.03.13.23287210

**Authors:** Farhad Ali Shah, Naila Shoaib, Asim Iqbal, Nazia Jamil, Rida Batool, Rimsha Munir, Ameem Lutfi, Nousheen Zaidi

## Abstract

The present study investigated the clinical and hematological effects of chronic lead exposure in the population residing in *Shadi Pura*, a small industrial zone in Lahore, Pakistan. A cross-sectional analysis of 149 participants recruited through health camps was conducted to explore the hematological manifestations of environmental lead exposure, focusing on various red blood cell (RBC) indices and morphology. Moreover, the study examined the differences in the impact of lead exposure on RBC indices and morphology between men, women, and children. Participants exhibited symptoms of lead poisoning, including fatigue, muscle pain, and headache, with a significant percentage of women (44%) reporting miscarriages. Iron deficiency anemia was highly prevalent among all sub-groups of the study population, with adult females showing a significantly higher prevalence than adult males. Male children were the most affected subgroup, with 93% displaying anemia. The RBC count in children remained unchanged, while 31% of male and 7% of female participants displayed elevated RBC counts. RBC indices, mainly mean corpuscular volume (MCV) and mean corpuscular hemoglobin (MCH), were below normal levels, with children being more affected than adults and adult males being the least affected group. Furthermore, RBC morphology was severely affected, with a considerable proportion of females and children displaying hypochromic microcytic morphology. Our results highlight variations in the hematological impacts of lead exposure in different gender and age cohorts. Overall our findings underscore the urgency of addressing the issue of environmental lead exposure in similar industrial zones. It is critical to implement appropriate measures to reduce lead exposure and enhance the infrastructure for safe drinking water and waste disposal to protect the health of populations in such areas.

## Introduction

Lead is a cumulative toxicant that poses a significant threat to human health. Children are especially susceptible to the toxic effects of lead, and it may cause permanent and irreversible damage to their developing brains and nervous systems [1]. Even low levels of lead may contribute to behavioral problems, learning deficits, and lowered IQ in young children. Lead also causes long-term harm in adults, including an increased risk of high blood pressure and kidney damage. Exposure of pregnant women to high levels of lead can cause miscarriage, stillbirth, premature birth, and low birth weight. Long-term lead exposure may cause anemia and hypertension in adults [1].

According to a recent systematic review conducted by *Ericson et al*. [2], Pakistan was found to have the second-highest mean childhood blood lead levels (BLLs) among low- and middle-income countries (LMICs), with an estimated value of 9.27 μg/dL (**Figure 1** [3]). The review further estimated that approximately 46.7 million children in Pakistan have BLLs exceeding the recommended threshold of 5 μg/dL [2]. These findings underscore the urgent need for effective policies and interventions to address the significant public health threat posed by lead exposure in Pakistani children.

**Figure 1:**
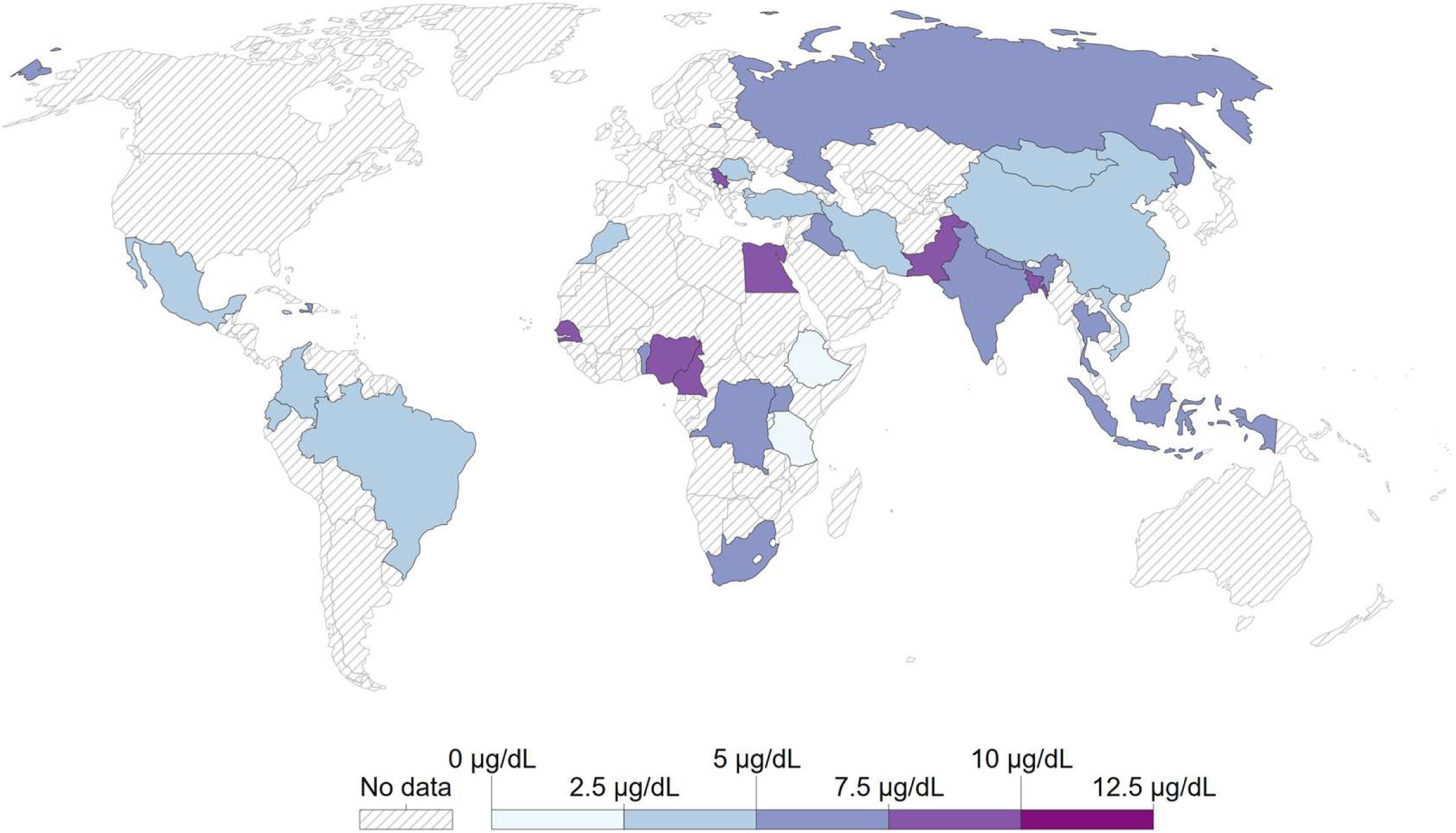
Mean childhood blood lead levels (BLLs) among low- and middle-income countries. Data from Ericson *et al*. (2021) via OurWorldInData.org/lead-pollution.

It is widely reported that increased levels of lead in the bloodstream are associated with a decrease in the body’s iron stores, thereby increasing the risk of anemia. *Słota et al*. [4] systematically reviewed the literature to study the association between lead poisoning and the body’s constitutive iron status. The systematic analysis included a comprehensive review of various epidemiological studies that examined several iron status markers, including hemoglobin, ferritin, serum iron, and total iron binding capacity. However, the results of studies that used hemoglobin, serum iron, or ferritin as iron status markers displayed some inconsistencies, with some studies indicating a negative correlation between lead-exposure markers and body iron status [4], while others showed no significant correlation, and some studies demonstrated an uncertain association. Interestingly, studies that utilized mean corpuscular volume (MCV) or red cell distribution width (RDW) as markers of iron status consistently showed a negative correlation between lead poisoning and iron status, albeit these studies were less frequent [5-8].

Another recent study indicated that blood lead concentrations were associated with increased odds of high RDW, and these associations were more pronounced in women and those with low-to-normal MCV [9]. Moreover, lead poisoning patients have been frequently shown to display hypochromic microcytic anemia –a type of anemia in which the red blood cells (RBCs) are smaller and paler than normal, with a reduced amount of hemoglobin [10]. These findings indicate that lead toxicity induces specific changes in red blood cells’ size, shape, and quality. Nevertheless, a scarcity of comprehensive investigations exists regarding the effects of lead poisoning on the morphology of red blood cells, with some previous studies producing contradictory results.

This study investigates the hematological effects of chronic lead exposure on a population residing in *Shadi Pura*, a small locality in Lahore, Pakistan. This area has numerous iron foundries crammed in a radius of ~20km. These iron foundries melt scrap, ingot, and other forms of iron and pour the resulting molten metal into molds to produce shaped products. This area’s soil and water samples contain hazardous lead levels. Here, we studied the hematological manifestations of environmental lead exposure in the area residents with an emphasis on RBC count, morphology, and indices. Furthermore, we evaluated whether the effects of lead exposure on hematological parameters differed between men, women, and children.

Overall, the study highlights the detrimental effects of chronic lead exposure on the health of individuals residing in areas with high concentrations of industrial activity. The results of this study have important implications for policymakers, public health professionals, and industries, as they underscore the need to implement measures to mitigate the negative impacts of environmental pollutants on human health.

## Materials and Methods

### Study Design

The study aimed to investigate the prevalence of anemia among residents of *Shadi Pura* (**Supplementary Figure 1**), a small residential locality (Population: ~51195) in Lahore, Pakistan, and to evaluate the association between environmental lead toxicity and hematological health. The study area was selected based on the presence of numerous metal foundries within a radius of ~20km, which melt scrap, ingot, and other forms of iron, and pour the resulting molten metal into molds to produce shaped products.

Data collection involved two main components: environmental lead toxicity assessment and hematological health assessment (**Supplementary Figure 2**). Environmental lead toxicity was assessed by collecting soil and water samples from different locations in *Shadi Pura*, including those in close proximity to the iron foundries. Hematological health was assessed through a blood test to determine the prevalence of anemia among study participants.

### Collection and Analysis of Environmental Samples

In order to investigate the quality of the tap water and soil in this area, samples were collected on December 13, 2021. A total of 0.7L of cold water samples were collected from the kitchen faucets of various *Shadi Pura* households, while 1 kg of composite soil samples was collected from locations identified as frequently visited areas by the study participants. All samples were collected in labeled sterilized flasks and were maintained in a refrigerated environment (4°C) during transportation to maintain the cold chain.

The water samples were sent to the *Soil and Water Testing Laboratory for Research*, Agriculture Department, Government of Punjab, Pakistan, where the water samples were tested for mineral elements and heavy metals, including Lead (Pb), Copper (Cu), Zinc (Zn), and Nickel (Ni). Standard procedures outlined in the United States Department of Agriculture (USDA) Handbook and atomic absorption were employed for the assessment [11].

The soil samples were sent to Central Labs at Syed Babar Ali School of Engineering Sciences, LUMS University, for Scanning Electron Microscopy with Energy Dispersive X-Ray (SEM-EDX) analysis. The soil samples were ground to a fine powder using an agate mortar and pestle and analyzed using FEI Nova NanoSEM 450 (FEI, Oregon).

Water and soil samples were also screened for heavy metal-resistant bacteria. Specifically, selected heavy metals, including Lead (Pb), Copper (Cu), Zinc (Zn), and Nickel (Ni), were added to standard Luria Bertani (LB, Peptone 10.00 g/L, yeast extract, 5.00 g/L, NaCl 5.00 g/L, and agar 30.00 g/L. pH:7.00) agar plates. Two concentrations of heavy metals (500 µg/mL and 100 µg/mL) were used for the screening protocols. Colony Forming Units (CFUs)/mL were determined using the standard pour plate technique, and plates were monitored after 24 hours of incubation at 37°C.

### Participant recruitment, ethics, and blood sample collection

The current study was conducted following the Helsinki Declaration and was approved by the Cancer Research Center Ethics Committee at University of the Punjab. Participants were recruited through health camps organized in *Shadi Pura*, which were arranged in collaboration with the *Haqooq-e-Khalq Movement* (HKM), a group of political activists who focus on projects related to public health and climate justice. Prior to the health camps, a door-to-door campaign was conducted to encourage residents of the area to attend.

A total of 149 participants attended the health camps, which were organized in collaboration with HKM and Hormone Lab, Pakistan. Of these participants, 68 were less than 15 years of age, while the remaining were above 15. Before sample and data collection, informed consent was obtained from each participant, and for minors, consent was obtained from their guardians.

Intravenous blood samples were collected from all participants using vials containing an EDTA-anticoagulant agent (BD Biosciences) and Clot Activator Tubes. Samples were immediately transported to Hormone Lab Lahore for processing, which included analysis of complete blood counts (CBC) and determination of serum ferritin levels. Additionally, information on symptoms, clinical history, and demographics was collected for each participant at the time of sample collection.

### Complete blood counts (CBC) and examination of the peripheral blood smears

For complete blood counts, EDTA samples were acquired on the multiparameter automated hematology analyzer (Swelab Alfa Plus system) according to the manufacturers’ guidelines. For morphology identification of blood cells, Giemsa (Merck) staining was performed on peripheral blood smears according to the manufacturer guidelines. Samples were imaged with a light microscope (Olympus CX31) at 40x and oil immersion (100x) magnification. The abnormality status of various parameters was determined using the standard values for each of these parameters.

### Determination of serum ferritin levels

Serum ferritin levels were determined with the *in vitro* chemiluminescence immunoassay (CLIA) kit by using the MAGLUMI 800 chemiluminescence immunoassay analyzer according to the manufacturer’s guidelines. The abnormality status of serum ferritin levels was determined by comparing the results with standard serum ferritin levels.

### Determination of Blood Lead levels

In order to ascertain the blood lead levels, venous blood specimens were acquired using EDTA vials and subsequently dispatched to Chugtai Lab, a commercial laboratory.

### Statistical Analysis

Data was analysed via GraphPad Prism 9.0.0 and MS-Excel 2016 using descriptive statistics. Continuous variables were presented as mean ± SD, while categorical variables were described as frequencies or percentages. The differences between groups were analysed by ANOVA or t-test (paired or unpaired), where applicable.

## Results

### Lead Contamination in Environmental Samples

We surveyed the drinking water sources of residents in *Shadi Pura* and found that 90% consumed tap water as their primary source. In order to determine the presence of heavy metal-resistant bacteria in the tap water of Shadi Pura, water samples were collected and screened. The results indicated that the bacteria present in the tap water were resistant to high concentrations (500 µg/mL) of various heavy metals, including Lead (Pb), Copper (Cu), Zinc (Zn), and Nickel (Ni) (**Supplementary Table 1**). Bacterial strains isolated from polluted environments are known to be tolerant to higher concentrations of metals than those isolated from unpolluted areas [11]. These results suggest potential lead contamination in tap water samples from Shadi Pura, which may pose health risks to the residents.

The concentration of the selected heavy metals in the water samples was determined through atomic absorption spectroscopy to confirm these findings further. The results indicated that the concentration of lead (Pb) (**Figure 2a, Supplementary Table 2**) was significantly higher than the permissible limit for drinking water set by the World Health Organization (WHO). However, the concentration of all other tested heavy metals was within the permissible limits set by the WHO. Other physicochemical parameters of the water sample were also analyzed, and the results are presented in **Supplementary Table 2**, along with the recommended limits set by the WHO and the Government of Punjab, Pakistan. The electrical conductivity, pH, and concentration of soluble salts were within the standard range.

**Figure 2:**
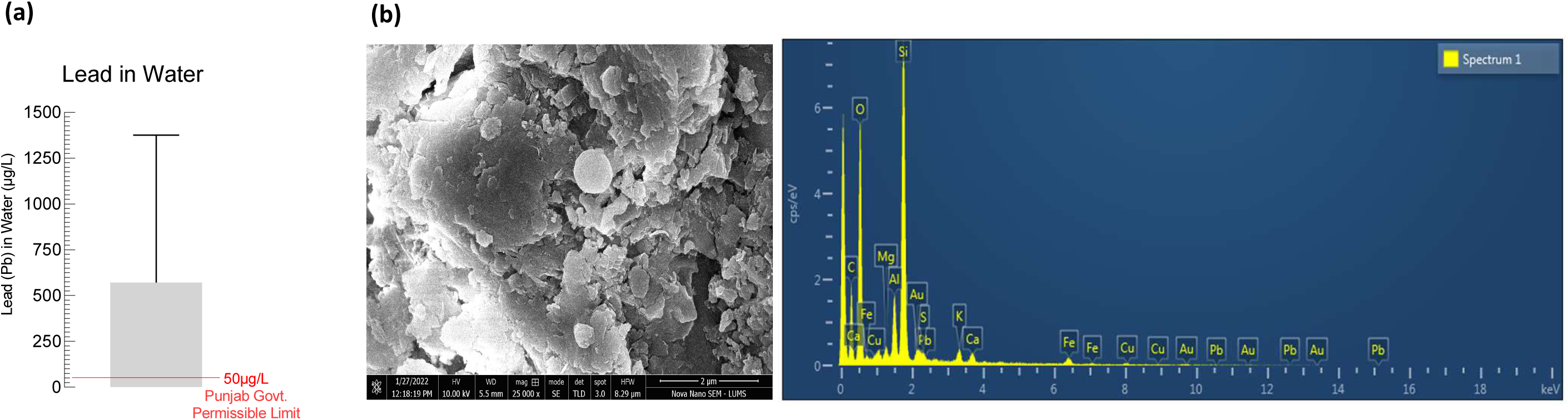
Lead contamination in water and soil samples from the study area. **(a)** *Bar graph* showing the lead concentration in the tap water sample from the study area. **(b)** SEM-EDX maps and point analysis spectra of a soil sample from the study area.

Next, we screened soil samples from Shadi Pura to identify heavy metal-resistant bacteria. The results showed that the bacteria present in the soil samples also exhibited resistance to high concentrations (500 µg/mL) of various heavy metals, including Lead, Nickel, Iron, Zinc, Copper, and Chromium (**Supplementary Table 3**).

Additionally, high-resolution scanning electron microscopy (SEM) images of the soil sample surface topography were obtained, and the SEM-EDX spectrum of the soil sample was analyzed. The SEM-EDX analysis revealed the presence of Si, O, C, Mg, Al, Cu, Pb, S, K, Au, and Fe (**Figure 2b**). Notably, the EDX spectrum showed low-intensity peaks for lead, despite its concentration being as high as 2.29% (weight %) (**Supplementary Table 4**) of the sample. This finding is consistent with previous research, suggesting that Pb particles may form physical aggregates with other soil constituents, making it difficult to detect the Pb peaks using SEM-EDX analysis [12].

### Symptoms of Lead Poisoning and Blood lead levels

We conducted a structured interview to collect data on demographic factors and common symptoms of lead poisoning among adult patients attending health camps organized for the residents of *Shadi Pura*. **Supplementary Table 5** presents information on the demographic characteristics of the study population, while **Figure 3a** displays the percentage of participants who reported various lead poisoning symptoms. The most commonly observed symptoms among the study participants were fatigue (91%), muscle pain (76%), and headache (73%). The participants also reported severe manifestations for a few of these symptoms, with severe fatigue being the most commonly reported **Figure 3b**). In addition, 44% of the women reported miscarriges (**Figure 3b**), while 13% reported premature births. We analyzed the blood lead levels of the study participants to confirm elevated blood lead levels and observed an average blood lead level of 16µg/dL± 2.9 (**Figure 3c**).

**Figure 3:**
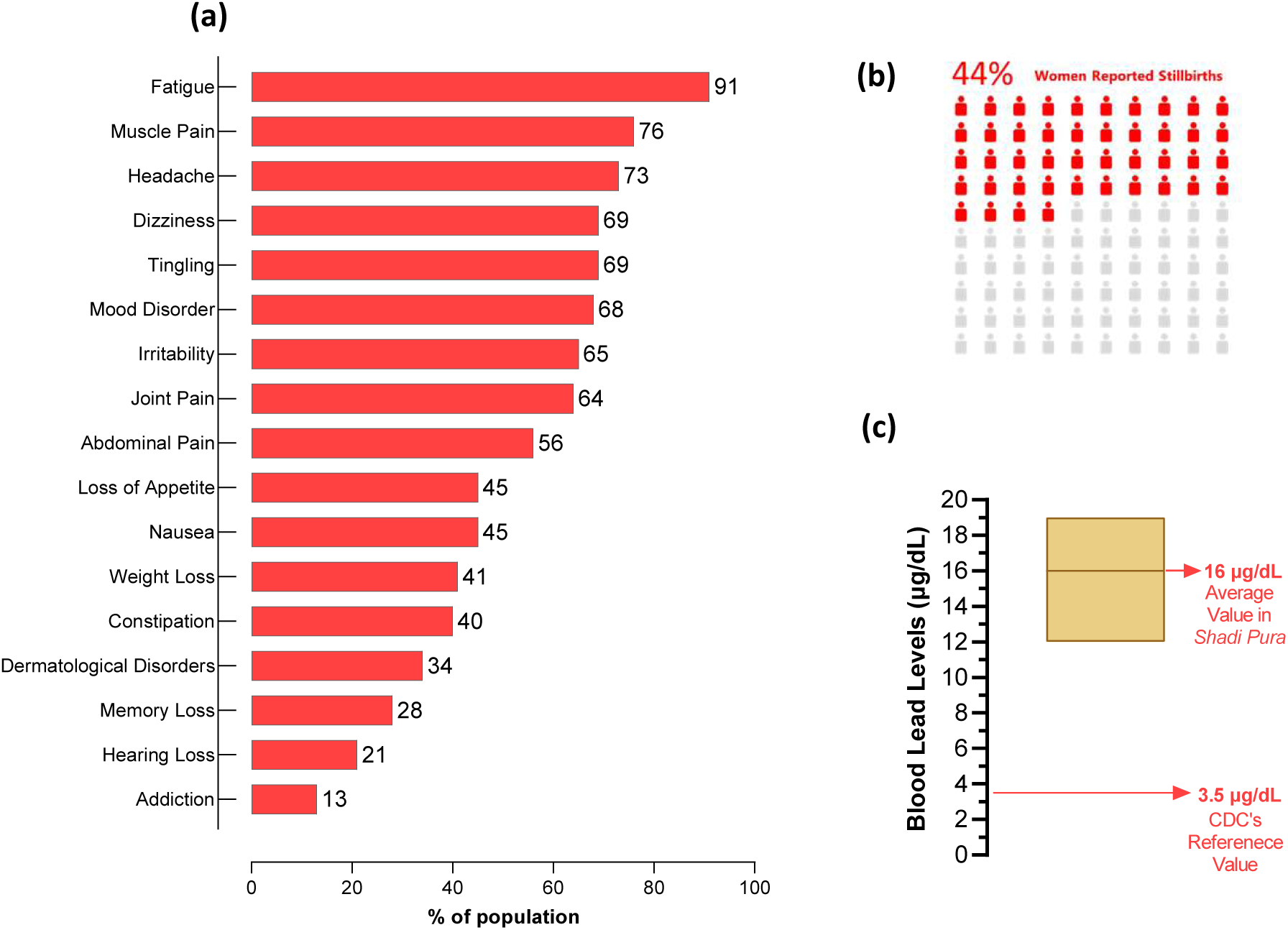
Symptoms of lead poisoning and blood lead levels in the study population. **(a)** *Bar graph* showing the percentage of the population displaying various symptoms of lead poisoning. **(b)** *Infographic* displaying the percentage of women who self-reported miscarriages. **(c)** *Box-plot* displaying the average blood lead levels in the area residents and CDC current reference value.

### Prevalence of Anemia

Subsequently, the present study aimed to investigate the incidence of iron deficiency anemia in the lead-exposed population of *Shadi Pura* **(Figure 4a, Table 1**). The investigation revealed a considerable prevalence of low hemoglobin levels across all sub-groups of the study population. Specifically, the adult female cohort (55%) displayed a significantly higher prevalence of low hemoglobin levels in comparison to their male counterparts (42%) **(Figure 4a, Table 1**).

**Table 1:** Haematological Indices and iron status of the Study Population. Trends were marked if the abnormality was observed in at least 5% of the population. (Trend Key: ↓;decreased, ↑;increased, ↔;unchanged)

|  |  |  |  |  |  |  |
| --- | --- | --- | --- | --- | --- | --- |
| Parameter | Study Group | Reference Values | Observed Values (Mean ± S.D) | Range | Trend observed | % Abnormality |
| **Haemoglobin (Gm/dl)** | Female | 12-16 | 11.53±1.62 | 6.3-14.5 | ↓ | 55 |
|  | Male | 14-18 | 13.98±1.66 | 9.8-16.1 | ↓ | 42 |
|  | Children (f) | 12-16 | 11.49±1.59 | 6.5-13.7 | ↓ | 53 |
|  | Children (m) | 14-18 | 11.96±1.49 | 7.6-14.2 | ↓ | 93 |
| **RBC (Mil/cmm)** | Female | 4.0-5.5 | 4.67±0.44 | 3.7-6.1 | ↑ | 7 |
|  | Male | 4.0-5.5 | 5.21±0.45 | 4.3-5.8 | ↑ | 31 |
|  | Children (f) | 4.0-5.5 | 4.64±0.31 | 4-5.7 | ↔ |  |
|  | Children (m) | 4.0-5.5 | 4.92±0.38 | 4.2-5.9 | ↔ |  |
| **Hematocrit (%)** | Female | 36-47% | 34.38±4.57 | 20-44 | ↓ | 56 |
|  | Male | 40-54% | 41.46±4.75 | 29-48 | ↓ | 35 |
|  | Children (f) | 36-47% | 33.28±4.37 | 19-39 | ↓ | 61 |
|  | Children (m) | 40-54% | 34.60±3.78 | 24-41 | ↓ | 93 |
| **MCV (f/L)** | Female | 77-95 | 73.60±8.01 | 47-88 | ↓ | 49 |
|  | Male | 77-95 | 79.27±5.98 | 64-91 | ↓ | 12 |
|  | Children (f) | 77-95 | 72.22±9.17 | 45-85 | ↓ | 61 |
|  | Children (m) | 77-95 | 70.50±8.50 | 49-80 | ↓ | 53 |
| **MCH (Pgm)** | Female | 27-32 | 25.05±3.31 | 15-30 | ↓ | 55 |
|  | Male | 27-32 | 27.27±2.47 | 20-31 | ↓ | 19 |
|  | Children (f) | 27-32 | 26.47±11.28 | 15-89 | ↓ | 67 |
|  | Children (m) | 27-32 | 24.13±3.50 | 15-29 | ↓ | 67 |
| **MCHC (G/L)** | Female | 31-35 | 33.53±1.27 | 30-35 | ↔ |  |
|  | Male | 31-35 | 33.92±1.23 | 31-36 | ↔ |  |
|  | Children (f) | 31-35 | 34.06±1.22 | 31-36 | ↔ |  |
|  | Children (m) | 31-35 | 34.23±1.38 | 31-38 | ↑ | 13 |
| **Ferritin** | Female | 13-232 | 34.53±30.36 | 3.9-120 | ↓ | 29 |
|  | Male | 25-350 | 114.82±84.57 | 9.1-347.3 | ↔ |  |
|  | Children (f) | 13-232 | 32.83±23.42 | 2.7-83.4 | ↓ | 23 |
|  | Children (m) | 25-350 | 41.45±26.36 | 13.6-67.4 | ↓ | 50 |

**Figure 4:**
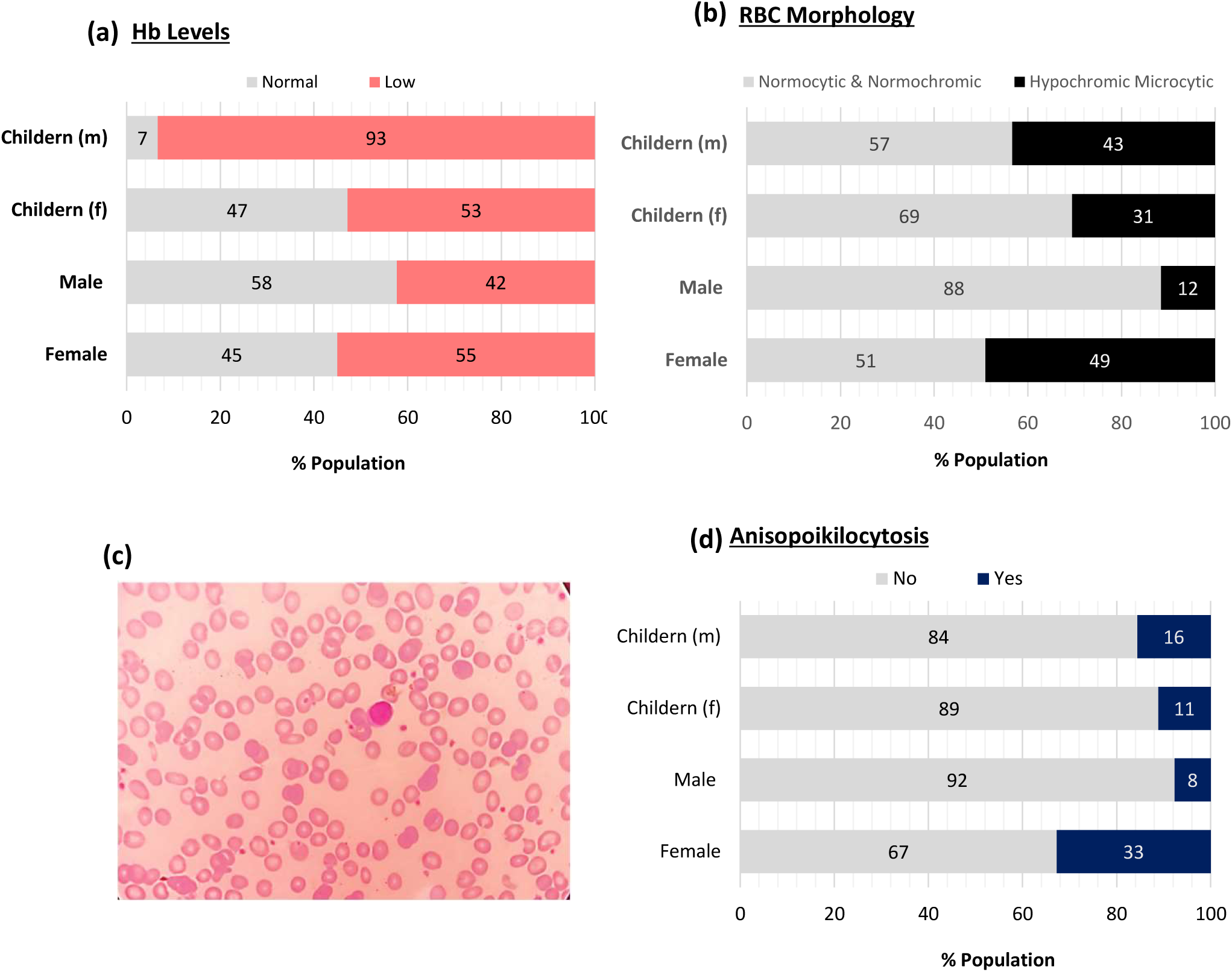
Prevalence of anemia and RBC morphology in the study population. Percentage of the different population sub-groups displaying regular or irregular **(a)** hemoglobin levels and **(b)** RBC morphology. (c) Hypochromic microcytic morphology observed in a representative sample from the study area. (d) Percentage of the different population sub-groups displaying anisopoikilocytosis.

On the other hand, male children exhibited a higher incidence of low hemoglobin levels than their female peers. Nonetheless, this observation may be attributed to the lower median age of the former group. Notably, studies on the correlation between iron deficiency and elevated blood lead levels have established their utmost significance in young children aged between 1 to 2 years, with weaker associations in older adolescents [4].

### Red Blood Cells Count and Morphology

Next, the impact of environmental lead exposure on red blood cell (RBC) count and morphology was investigated. The study revealed that 7% of female participants showed an increase in RBC counts, while a significantly larger proportion of male participants, i.e., 31%, exhibited elevated RBC counts (**Table 1**). In contrast, male and female children displayed a comparable rise of 3% in RBC counts.

Furthermore, hematocrit, which measures the volume percentage of RBCs in the blood, was significantly lower in all population subgroups, with male children (93%) being the most affected group. This indicates a decrease in the amount of space occupied by RBCs in the blood.

The study also found that RBC morphology was severely impacted in the lead-exposed population, with a significantly large population displaying hypochromic microcytic morphology (**Figure 4b-c**). For the female population, the percentage of participants displaying hypochromic microcytic morphology (49%) was the highest, followed by male children (43%) and female children (31%). In the adult male population, this percentage was 12%.

The RBC indices mean corpuscular volume (MCV) and mean corpuscular hemoglobin (MCH) were also significantly reduced in our population, with children being the most affected subgroup (**Table 1**). Conversely, mean corpuscular hemoglobin concentration (MCHC) was the least affected RBC index (**Table 1**). Additionally, the study observed that 33% of the adult female population exhibited anisopoikilocytosis (**Figure 4d**).

We also examined the levels of blood ferritin and observed that a considerable proportion of female participants (29%) displayed ferritin deficiency, whereas the ferritin levels in the male participants were found to be generally normal, as indicated in **Table 1**. In addition, both male and female children displayed lower levels of ferritin, with a significantly greater proportion of male children (50%) presenting with values lower than the normal range.

## Discussion

The historical use of tetraethyl lead in petrol as an anti-knocking agent dispersed at least 9 million tonnes of lead into the environment [13]. The global phase-out of leaded gasoline began in the 1970s and was completed in 2021 when Algeria became the last country to ban it. This phase-out has had a significant impact on the average blood lead levels (BLL) in people in high-income countries, where it has decreased significantly. For instance, studies have reported that the BLL in people aged 1-74 in the USA decreased by 78% between 1976 and 1991, from 12.8 μg/dL to 2.8 μg/dL [14].

Despite a notable decrease in BLL in high-income countries, recent research indicates that low- and middle-income countries still experience significantly elevated levels of BLL [2]. Among these countries, Pakistan stands out with some of the highest BLLs in both the adult and pediatric populations. The average BLL in adults is reported to be 11.36±5.20 μg/dL, while in children, it is 9.27±3.17 μg/dL [2].

The sources of lead exposure in low-income countries are distinct from those in high-income countries. After the phase-out of leaded petrol, the major sources of high lead exposure are lead-based enamel paints or lead pipes in residential settings in countries such as the USA. However, in low-income countries, critical sources of lead exposure include informal lead acid battery recycling and manufacture, metal mining and processing, electronic waste, and the use of lead as a food adulterant, primarily in spices [2]. Additionally, many low- and middle-income countries lack proper regulations on industrial activities that produce lead emissions, and lead-containing consumer products are still prevalent in some areas. In Pakistan, especially in the residential areas close to industrial areas, excess lead in drinking water is identified as a significant source of lead toxicity [15]. Industries that use lead in their production processes can contaminate the environment through untreated, toxic effluent that is sometimes released directly into nearby water systems or dumped on the ground, contaminating soil and groundwater.

Our data show a high level of lead contamination in water and soil samples collected from *Shadi Pura*, a prominent industrial zone in Lahore, Pakistan’s second-largest city. The presence of heavy metal-resistant bacteria in the water sample suggests widespread contamination. People living in the residential quarters of this neighborhood are chronically exposed to lead; as a result, they display very high levels of BLL and common symptoms of lead poisoning. This study was focused on studying the hematological health of this lead-exposed population.

The hematological system is generally regarded as a sensitive target tissue of lead exposure [4]. Here we investigated the incidence of iron deficiency anemia in the lead-exposed population of *Shadi Pura*.

Our findings indicate a considerable prevalence of low hemoglobin levels across all sub-groups of the study population. The adult female cohort displayed a significantly higher prevalence of low hemoglobin levels compared to their male counterparts. This observation is consistent with previous studies indicating a higher incidence of anemia in adult women due to factors such as menstrual blood loss, pregnancy, and lactation [16, 17]. However, it is worth noting that the prevalence of anemia among men was still relatively high.

Interestingly, male children exhibited a higher incidence of low hemoglobin levels than their female peers, which may be attributed to their younger age. Studies have shown that iron deficiency and elevated blood lead levels have the strongest correlation in young children aged between 1 to 2 years, with weaker associations in older adolescents [4]. The high incidence of low hemoglobin levels among male children in *Shadi Pura* may be indicative of the need for further investigation and intervention to prevent and manage iron deficiency anemia in this population

Based on our data, the lead-exposed population displayed an elevated RBC count, with the most notable effect observed among males, as indicated by both the proportion of affected individuals, which accounted for 31% of the male population and the mean RBC count. Previous studies have also indicated that exposure to lead can increase the number of red blood cells (RBCs) in the body, leading to a higher RBC count [5, 6]. The etiology of high red blood cell (RBC) count resulting from lead exposure remains incompletely understood; however, it is hypothesized to involve erythropoietin (EPO) production stimulation, the hormone responsible for RBC production in the bone marrow. Lead may impede the functioning of enzymes involved in heme synthesis, which is essential for hemoglobin production. Consequently, lead exposure can cause anemia, triggering EPO production to stimulate RBC production. Additionally, it has been suggested that the rise in RBC count may be due to diminished erythrocyte function rather than a decrease in RBC count, leading to the stimulation of RBC formation [6]. Previous research has reported sex-based disparities in the correlation between lead toxicity and red blood cell (RBC) count among school-aged children. Specifically, the findings indicate that boys are more susceptible to surpassing the recommended RBC count threshold than girls [6]. This disparity may be attributed to higher lead exposure among males than females. However, further research is required to understand the difference in male and female

One of the main objectives of this study was to evaluate the influence of lead on red blood cell morphology and to investigate how this interaction is modulated by patient age and sex. We found that lead exposure severely impacts RBC morphology, with a significantly large proportion of the population displaying hypochromic microcytic morphology. The percentage of participants with hypochromic microcytic morphology was the highest in adult females, followed by male and female children. Previous studies have shown that lead exposure can affect RBC morphology through various mechanisms, including inhibition of delta-aminolevulinate dehydratase [18], oxidative stress [19], and disruption of the RBC membrane structure [20]. As observed in the present study, these effects can result in hypochromic microcytic morphology and anisopoikilocytosis.

In conclusion, our study highlights the significant impact of lead exposure on the hematological system, particularly regarding iron deficiency anemia and red blood cell morphology. Our findings suggest that individuals in the lead-exposed population of *Shadi Pura* are at an increased risk of anemia, particularly women and children. Additionally, lead exposure appears to increase the RBC count, with the most significant effect observed among males. Furthermore, lead exposure severely impacts RBC morphology, with a large proportion of the population displaying hypochromic microcytic morphology. These findings underscore the importance of continued efforts to mitigate lead exposure and its effects on the hematological system. Developing and implementing effective interventions to prevent and manage iron deficiency anemia and other hematological disorders in populations at risk of lead exposure is crucial. Further research is also necessary to better understand the mechanisms underlying the observed effects and identify new prevention and treatment strategies.

## Data Availability

All data produced in the present study are available upon reasonable request to the authors

---

**Supplementary Figure 1:**
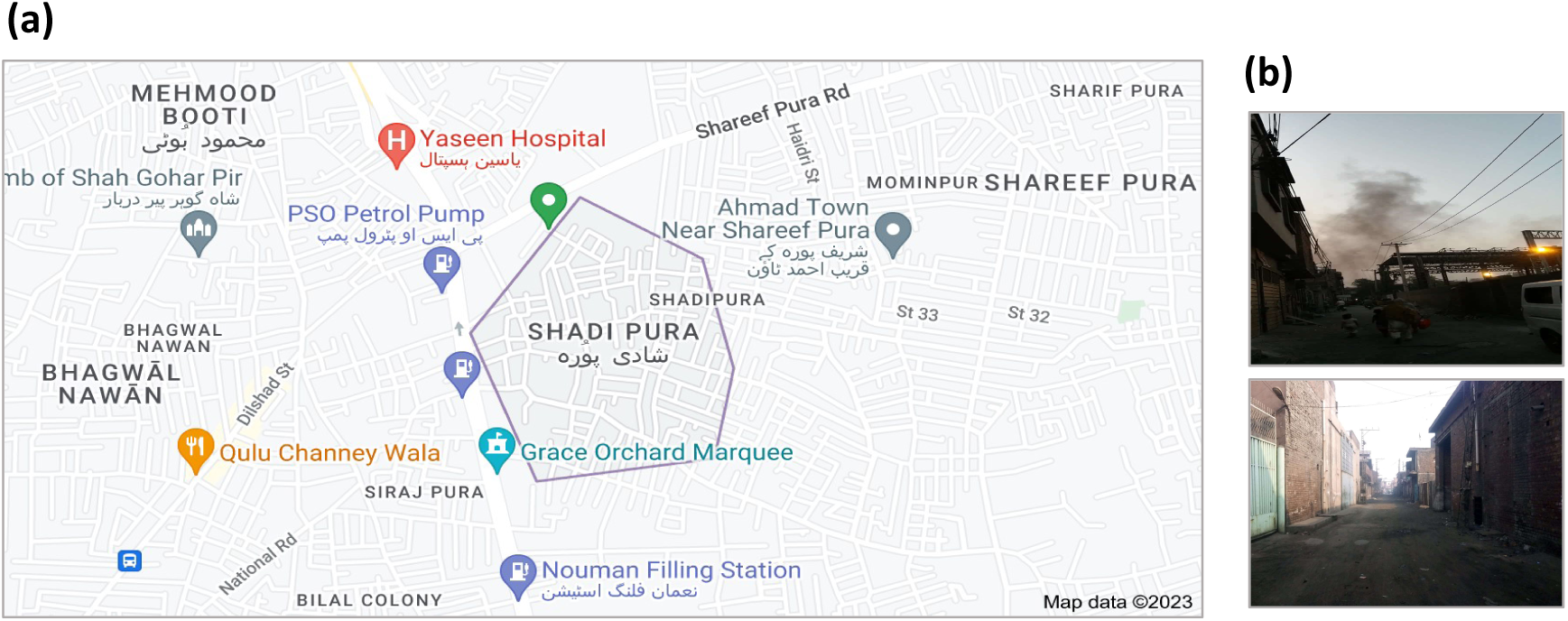
Google map and images from the *Shadi Pura*. *Shadi Pura* a small residential locality (Population: ~51195) in Lahore, Pakistan. **(a)** *Google map* of Shadi Pura **(b)** Images from *Shadi Pura*.

**Supplementary Figure 2:**
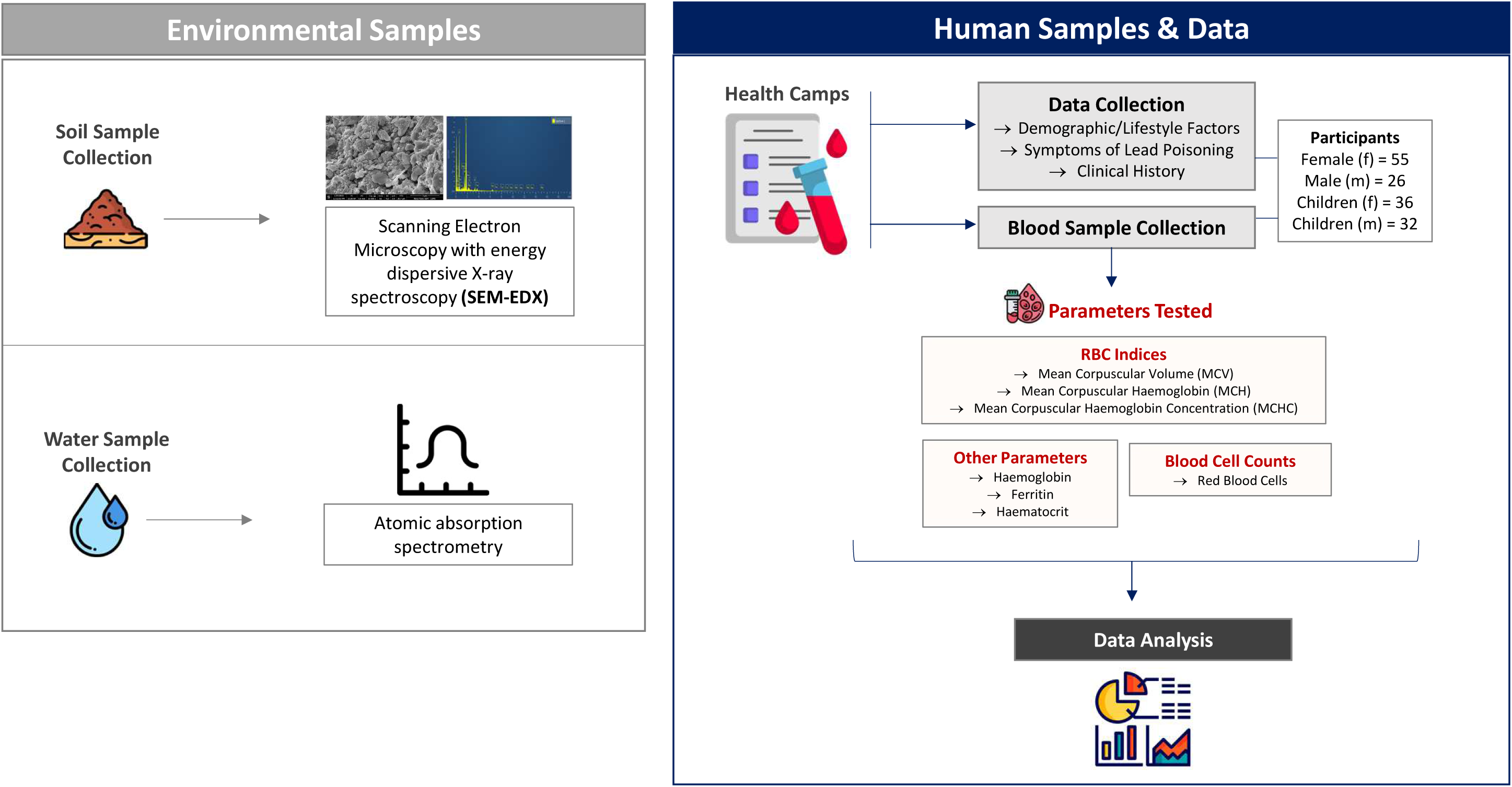
Study Design. Data collection involved two main components: environmental lead toxicity assessment and hematological health assessment.

**Supplementary Table 1:** Heavy–metal resistant bacteria in water samples.

|  |  |  |  |
| --- | --- | --- | --- |
| **Metal** | **Conc. Tested**<br>**(µg/mL)** | **Bacterial Resistance** | **CFU/mL** |
| Lead | 500 | Yes | 1000 |
| Copper | 500 | Yes | 20000 |
| Zinc | 500 | Yes | 1700 |
| Nickel | 500 | Yes | 2000 |

**Supplementary Table 2:** Physiochemical properties of a selected water sample from *Shadi Pura*.

| Parameter | Observed Value | Punjab Govt. limits | WHO limits [1] |
| --- | --- | --- | --- |
| pH | 7.00 | 6.5–8.5 | 6.5–8.5 |
| Electrical conductivity ($\mu\text{S}/\text{cm}$) | 295 | NA | $\leq 400$ |
| Sodium ($\text{mg}/\text{L}$) | 6.18 | NA | $\leq 20$ |
| Calcium ($\text{mg}/\text{L}$) | 53 | $\leq 500$ | $\leq 60$ |
| Chloride ($\text{mg}/\text{L}$) | 18 | $\leq 250$ | $\leq 250$ |
| Carbonate ($\text{mg}/\text{L}$) | 9.76 | $\leq 500$ | $\leq 60$ |
| Bicarbonate ($\text{mg}/\text{L}$) | 119 | NA | NA |
| Residual Sodium Carbonate (emp) | ND | NA | NA |
| Sodium Adsorption Ratio | 0.23 | NA | NA |
| **Heavy Metals** |  |  |  |
| Lead (Pb) ($\mu\text{g}/\text{L}$) | 1550 | $\leq 50$ | $\leq 10$ |
| Nickel (Ni) ($\mu\text{g}/\text{L}$) | ND | $\leq 20$ | $\leq 20$ |
| Copper (Cu) ($\mu\text{g}/\text{L}$) | 570 | $\leq 2000$ | $\leq 2000$ |
| Zinc (Zn) ($\mu\text{g}/\text{L}$) | 580 | $\leq 5000$ | $\leq 3000$ |

**Supplementary Table 3:** Heavy–metal resistant bacteria in soil samples.

| **Supplementary Table 3:** Heavy-metal resistant bacteria in soil samples. |  |  |  |
| --- | --- | --- | --- |
| **Metal** | **Conc. Tested**<br>**(µg/mL)** | **Bacterial Resistance** | **CFU/mL** |
| Lead | 500 | Yes | 440 |
| Copper | 500 | Yes | 20 |
| Zinc | 500 | Yes | 1820 |
| Nickel | 500 | Yes | 540 |

**Supplementary Table 4:** EDX analysis of the soil sample.

| Supplementary Table 4: EDX analysis of the soil sample. |  |  |  |
| --- | --- | --- | --- |
| Sr. No. | Element | Weight % | Standard Label |
| 1 | C | 36.58 | C Vit |
| 2 | O | 39.5 | SiO<sub>2</sub> |
| 3 | Mg | 0.55 | MgO |
| 4 | Al | 2.76 | Al<sub>2</sub>O<sub>3</sub> |
| 5 | Si | 13.79 | SiO<sub>2</sub> |
| 6 | S | 0.43 | FeS<sub>2</sub> |
| 7 | K | 1.06 | KBr |
| 8 | Ca | 0.89 | Wollastonite |
| 9 | Fe | 2.14 | Fe |
| 10 | Cu | 0 | Cu |
| 11 | Pb | 2.29 | PbTe |
| 12 | Au | 0 | Au |

**Supplementary Table 5:** Population Demographics & Baseline Data.

| **Supplementary Table 5: Population Demographics & Baseline Data.** |  |  |
| --- | --- | --- |
| **Variable** | **Adults** | **Children** |
| **Age (Yrs., Mean ± SD)** |  |  |
| Male | 41.19 ± 21.73 | 6.9 ± 3.91 |
| Female | 38.63 ± 15.13 | 8.45 ± 4.61 |
| **Age (Yrs., Median)** |  |  |
| Male | 34 | 7.5 |
| Female | 37.5 | 9 |
| **Marital Status (%)** |  |  |
| Male | 73.08 | NA |
| Female | 81.82 | NA |
| **Number of Children (Median)** |  |  |
| Male | 4 | NA |
| Female | 4 | NA |
| **Weight (Kgs., Mean ± SD)** |  |  |
| Male | 65.81±11.03 | 20.53±8.25 |
| Female | 63.90±16.20 | 24.72±12.87 |
| **Height (inches., Mean ± SD)** |  |  |
| Male | 61.78±8.92 | 35.85±16.32 |
| Female | 59.90±2.99 | 29.65±21.78 |
| **Oxygen saturation (%, Mean ± SD)** |  |  |
| Male | 96.77±6.54 | 77.5±21.92 |
| Female | 96.75±5.82 | 92.5±8.48 |
| **Pulse Rate (bpm., Mean ± SD)** |  |  |
| Male | 86.31±14.62 | 60±14.14 |
| Female | 90.48±16.02 | 87.9±20.43 |
| **Blood Pressure (mmHg., Mean ± SD)** |  |  |
| **Systolic** |  |  |
| Male | 142.17±23.51 | NA |
| Female | 146.29±22.71 | NA |
| **Diastolic** |  |  |
| Male | 89.75±12.07 | NA |
| Female | 93.13±18.84 | NA |

## Notes

### Competing Interest Statement

The authors have declared no competing interest.

### Funding Statement

This study was funded by University of the Punjab.

### Author Declarations

The current study was conducted following the Helsinki Declaration and was approved by the Cancer Research Center Ethics Committee at University of the Punjab.

